# HIV TREATMENT OUTCOMES IN RURAL GEORGIA USING TELEMEDICINE

**DOI:** 10.1101/2020.11.02.20224600

**Authors:** Folake J. Lawal, Moshood O. Omotayo, Tae Jin Lee, Arni S.R. Srinivasan Rao, Jose A. Vazquez

## Abstract

**Background:** The dearth of specialized healthcare services contributes to the ongoing HIV epidemic. Telemedicine (TM) is a potential tool to improve HIV care, but little is known about its effectiveness when compared to traditional (face-to-face) (F2F) care in rural populations. The objective of this study is to examine the effectiveness of HIV care delivered through TM compared to F2F care.

**Methods:** We conducted a retrospective chart review of a subset of HIV patients who attended TM clinic in Dublin Georgia, and conventional F2F clinic in Augusta, Georgia between May 2017 to April 2018. All TM patients were matched to F2F patients based on gender, age, and race. HIV Viral Load (VL) and gain in CD4 counts were compared using T-test and Snedecor Statistics.

**Results:** 385 patients were included in the analyses (F2F=200, TM=185). Mean CD4 in the TM group was higher (643.9 cells/mm3) than the F2F group (596.3 cells/mm3) (p< 0.001). There was no statistically significant difference in VL reduction and control. Thirty-eight of eighty-five patients with detectable VL achieved viral suppression during the study period (F2F = 24/54, TM =14/31), with a mean change of −3.34 × 10^4^ and −1.24 × 10^4^ respectively, p = 1.00. Mean VL was F2F = 416.8 cp/ml, TM = 713.4 cp/ml, p = 0.3.

**Conclusion:** TM was associated with outcome measures comparable to F2F. Increased access to specialty HIV care through TM can facilitate HIV control in communities with limited healthcare access in rural US. Rigorous prospective evaluation of TM for HIV care effectiveness is warranted.

**Article Summary:** Telemedicine can be useful in improving access to specialist outpatient care for HIV and other chronic diseases, in remote communities with limited resources. Telemedicine can lead to similar outcomes when compared to traditional face-to-face outpatient consultations. This is especially true currently with COVID-19.

## Introduction

In the US, there are about 1,173,900 people above age 13, living with the Human Immunodeficiency Virus (HIV), with an estimated 13.8% of them undiagnosed (1). More than 86% of newly diagnosed persons were linked to care within 3 months of diagnosis in 2017. By the beginning of 2017, only 57.6% of newly diagnosed HIV patients were retained in care for the entire 2016 and only 61.5% of those retained achieved viral suppression (2). This rate of change is not on track to end AIDS by 2030 (3,4).

As with many chronic ailments, the challenges to HIV care and control are multifaceted. These challenges include social, geographical, health systems and economic factors (5). Access to health care by patients can be easily impacted by policies that translate to limited funding, limits to spending options or services covered as adjunct to health care such as transportation. The negative effects of location on healthcare utilization are well documented for persons in difficult-to-reach regions with a poor health infrastructure (6,7). A study reviewing 15 articles published between 1997 and 2010 reported multiple barriers to HIV care, with transportation being among the most commonly reported barriers (8). Multiple innovations have been employed to bridge the gaps such as using drones and empowering lower level providers with or without telemedicine support (9).

Geographical and transport constraints continue to play an important role in access to care across the US. This constraint plausibly has greater impact in the Southern states which have higher proportions of rural dwellers. Travel time to access health care has been shown to reduce the use of specialty care in rural dwelling veterans and impacted their health negatively (10,11). At our Augusta University facility, HIV patients commute up to 3-4-hour one way to receive the care they need. In 2018, the rate of new diagnoses in the southern states, was 18.4 per 100,000, about 5 percentage points higher than the national average of 13.3%. (1) The southern states accounted for 52% of new HIV cases nationwide. Furthermore, 23% of new HIV diagnoses were in rural and suburban areas in 2017 (12). Rural dwellers have inferior outcomes due to delays in diagnosis and linkage to care, and as well as poor retention and adherence rates are also recorded at higher rates in these areas (13–15). Of the barriers to care commonly reported, particularly in HIV patients, distance and transportation remains high on the list. Women living with HIV in the rural southeast US strongly endorsed transportation related issues with long distances as primary barriers to maintaining HIV care and appointments (16).

Telemedicine has enhanced ease of collaboration and support to improve health care access and delivery across the world (17). In the US, telemedicine has been shown to be effective to reduce travel distance, time, and cost of care in post vascular surgery follow up (18). More importantly, telemedicine has enabled global collaboration to close gaps in health care delivery. Resource poor countries have benefitted from specialty care with physician-to-physician connections or physician-to-patient connections to deliver essential health care to remote and rural dwellers (19). Countries with advanced health care have also demonstrated how telemedicine can reduce the burden of time on patients, and in some cases privacy for HIV patients (20). Effectiveness of telemedicine has been demonstrated in acute care to reduce the rates of urgent referral. It has also been effective for specialty consultation in ophthalmology cases managed by virtual consultation in Queensland and HIV/AIDS case consultations and discussions (21).

HIV care by infectious disease specialists and HIV trained “generalists” have been documented to deliver superior care when compared to non-HIV trained generalists in traditional outpatient settings (15,22). A study examining quality of care delivered by correctional facility physicians vs multidisciplinary infectious disease team delivered by Telemedicine in a prison population showed higher rates of virologic suppression in the telemedicine cohort (23). Using a physician - patient home care telemedicine model with stable and virally suppressed HIV patients, a randomized trial in Spain showed that 85% of participants endorsed savings with time and money. Additionally, there was a high (81%) acceptance rate and the clinical parameters were equivalent in both groups (20). Despite existing evidence of the potential value of telemedicine in improving access to specialty HIV care in underserved populations, there is a paucity of studies examining the effectiveness of TM in improving HIV clinical endpoints in rural US populations.

In this study, we examine effectiveness of telemedicine in delivery of direct long-term HIV care in rural Georgia. We conduct a head to head comparison with a parallel group of patients seen by the same group of Infectious disease (ID) physicians at a nearby tertiary institution using various parameters such as HIV PCR (Viral Load) (VL) and CD4 count.

## Methods

### Study population

The target population for this study comprises HIV patients in rural communities in South-East United States. The study population was drawn from two patient groups. The telemedicine (TM) group comprised all patients in the Dublin Department of Health HIV clinic database and the traditional group was drawn from the Augusta University (AU) (face-to-face) (F2F) HIV clinic patient database. All patients were seen by the same group of Infectious disease providers. The Dublin clinic is situated approximately three hours from Augusta University, in Augusta, GA and patients enrolled at Dublin clinic commuted from varying distances to the clinic, and are triaged and examined by the clinic nurse prior to video conference with an infectious diseases (ID) physician. Laboratory and necessary paperwork were transmitted securely to the AU ID physician prior to the visit. Documentation was done in the AU Electronic Medical Record, (EMR) and then transmitted to the Dublin clinic by fax.

### Study design

We conducted a retrospective data review for HIV patients enrolled and seen between May 2017 and April 2018 for both groups. Based on the preliminary TM pool data using unpaired simple effects analyses 250 subjects are needed to detect a statistical significance at p = 0.05, 2-tailed and 80% power of detection.

Inclusion criteria were age > 18 years and enrollment in care > 6 months. Exclusion criteria included pregnant patients, patients newly enrolled six months or less prior to May 2014. Charts with incomplete data were excluded. Data extracted included demographics, number of visits, dates of clinic attendance, CD4 cell count, VL, HIV resistance mutations, and major comorbidities.

The patients seen between that 12-month period included 1391 in the F2F arm and 236 for the TM arm. For the TM group 26 cases were enrolled for less than 6 months. Of the remaining 210, 25 had incomplete data leaving 185 as the TM study population.

The F2F sample pool of 1391 participants was stratified by gender, then race, followed by age, then a systematic sampling using 1:5 to 1:7 sampling interval from each stratified group to give a 263 cases, 31 were excluded for being enrolled less than 6 months. 232 left as study population, 32 had incomplete data leaving 200 included in the final study sample.

#### Definitions of variables

VL of <40 copies/ml were categorized as undetectable. Co-morbidities were extracted by International Classification of Diseases, Tenth revision, codes (ICD-10). New resistance was extracted from laboratory data in EMR and physician chart documentation of resistance mutations. Medication changes included: any change in medication class or formulation within review period as noted in physician notes, prescriptions, and medication list. This was categorized in to 5 groups, coded as: 0= No Change; 1= Newer medication/ physician preference; 2= Failure of therapy/ new resistance; 3= Adverse effect; 4= consolidate pill burden. Data extraction and coding was carried out by one investigator who is familiar with the electronic health system of the institutions.

#### Outcomes

The main outcome was rates of viral suppression and maintenance of suppression during the review period.

### Statistical analysis

The average CD4 count for each patient was calculated as the mean of the measurements from all visits recorded in the 12-month period for the patient. The average CD4 count for each group was computed as the mean of the average count for all patients in the group. t-tests were conducted to test differences in average CD4 count between the two groups (TM and F2F).

The difference between the first and last visit measurements were used to calculate the change in CD4 count for the patient. Based on the direction of change, patients were categorized into subgroups of those with increasing CD4 count and those without increasing CD4 count. The proportion of patients with increasing CD4 counts was calculated as number of patients with an increase in CD4 count divided by the total number of patients in the group. Using the chi-square test statistic, we performed a test of proportions to determine if there was a significant difference in the proportion of patients that showed an increase in CD4 counts in the F2F and TM populations. The t test was used to compare the mean increase in CD4 count between the two groups (TM and F2F), for the subgroup of patients with increasing CD4 count. The t-test was also used to compare the mean decrease in CD4 count between the two groups (TM and F2F) for the subgroup without increasing CD4 count.

The average VL for each patient was calculated as the mean of the measurements from all visits recorded in the 12-month period for the patient. The average VL for each group was computed as the mean of the average count for all patients in the group. T-tests were conducted to test differences in average VL between the two groups (TM and F2F).

The proportion of patients with decreasing VL was calculated as number of patients with decrease in VL divided by the total number of patients in the group. Using the chi-square test statistic, we performed a test of proportions to determine if there was a significant difference in the proportion of patients that showed decrease in VL in the F2F and TM populations.

The difference between the first and last visit measurements were used to calculate the change in VL for the patient. Based on the direction of change, patients were categorized into subgroups of those with decreasing CD4, VL and those without decreasing VL. The Snedecor Statistics was used to compare the mean decrease in VL between the two groups (TM and F2F) for the subgroup with decreasing VL. The Snedecor Statistics was also used to compare the mean increase in VL between the two groups (TM and F2F) for the subgroup without decreasing VL. For all tests, differences were considered statistically significant when p-value was less than 0.05 or the 95% confidence interval excluded the null value.

### Ethical approval

The study was approved by the Institutional Review Board at Augusta University and the Georgia Department of Public Health. A waiver of informed consent was granted by the Institution Review Board as no direct patient contact was planned, data gathered was coded and linked for reference, however no personal identifiers were disclosed.

## RESULTS

In this study, a total of 385 cases were included, (52.5% black, 82% females, F2F = 200, TM = 185). Distribution of comorbidities were similar between both groups with cardiovascular, renal, neuropsychiatric and diabetes being the most common as seen in Table 1.

**TABLE 1:** Key characteristics of face-to-face and Telemedicine HIV patients.

|  | **F2F** | **TM** |
| --- | --- | --- |
| **Gender** | N (%) | N (%) |
| Females | 106 (53) | 90 (48.6) |
| Males | 89 (44.5) | 95 (51.4) |
| Transgender | 5 (2.5) | 0 (0) |
| **Race** |  |  |
| Black | 164 (82) | 152 (82.2) |
| White | 30 (15) | 32 (17.3) |
| Others | 6 (3) | 1 (0.5) |
| **Age (years)** |  |  |
| 18-30 | 21 (10.5) | 18 (9.7) |
| 31-50 | 77 (38.5) | 68 (36.8) |
| 51+ | 102 (51) | 99 (53.5) |
| **Medication Change Code** |  |  |
| 0= No Change | 162 (81) | 158 (85.4) |
| 1= New therapy / Physician Preference | 19 (9.5) | 5 (2.7) |
| 2= Failure of Therapy | 8 (4) | 4 (2.2) |
| 3= Adverse Effect | 10 (5) | 10 (5.4) |
| 4= Consolidation for Pill Burden | 1 (0.5) | 8 (4.3) |
| **Resistance Code** |  |  |
| No New Resistance | 192 (96) | 183 (98.9) |
| New Resistance | 8 (4) | 2 (1.1) |
| **Comorbidities** |  |  |
| Cardiac | 117 (58.5) | 55 (29.7) |
| Respiratory | 17 (8.5) | 3 (1.6) |
| Diabetes | 26 (13) | 20 (10.8) |
| Renal disease | 63 (31.5) | 33 (17.8) |
| Neuropsychiatry | 75 (37.5) | 32 (17.3) |
| Liver | 17 (8.5) | 12 (6.5) |
| Dental | 11 (5.5) | 4 (2.2) |
| Alcoholism | 7 (3.5) | 5 (2.7) |
| Drugs | 16 (8) | 7 (3.8) |
| Cancer | 1 (0.5) | 4 (2.2) |
| Autoimmune Diseases | 2 (1) | 3 (1.6) |

We found the mean CD4 count in the TM group as shown in Table 2, was statistically higher (643.9 cells/mm3) than the F2F group (596.3 cells/mm3) (p<0.001). Among those with increased CD4 count, the increase in CD4 count was 120.76 cells/mm3 in the TM group and 134.52 cells/mm3 in the F2F subgroup (p =0.45).

**TABLE 2:** Clinical outcomes in face-to-face and Telemedicine HIV patients

|  | **F2F** | **TM** | **P-value** |
| --- | --- | --- | --- |
| Mean CD4 | 569.3 | 643.9 | <0.001<sup>a</sup> |
| Mean VL | 416.8 | 713.4 | 0.3<sup>a</sup> |
| Mean ΔCD4 | 134.52 | 120.76 | 0.45<sup>a</sup> |
| Mean ΔVL | -3.34 x 10<sup>4</sup> | -1.24 x 10<sup>4</sup> | 1<sup>b</sup> |
| VL Decrease | 0.44 | 0.45 | 1<sup>c</sup> |
| CD4 Increase | 0.41 | 0.43 | 1<sup>c</sup> |
| VL UD | 0.73 | 0.77 | 0.54<sup>c</sup> |
| **Abbreviations:** |  |  |  |
| F2F = face-2-face |  |  |  |
| TM = telemedicine |  |  |  |
| <sup>a</sup> = T-statistics |  |  |  |
| <sup>b</sup> = Snedecor statistics |  |  |  |
| <sup>c</sup> = Chi squared statistic |  |  |  |
| UD VL = Year-round Undetectable VL |  |  |  |

There was higher mean VL in the TM group (713.4 cp/ml), compared to the F2F group (416.8 cp/ml), but it was not statistically significant with p-value = 0.3 as shown in Table 2. Year-round rate of viral control was similar between both groups [(TM = 154/185 (77%), F2F= 146/200 (73%)]. There was no significant difference between the two populations regarding the viral suppression among patients who were not suppressed at the beginning of the study period. Thirty-eight of eighty-five patients with detectable VL achieved viral suppression before the end of the study period (F2F= 24/54, TM =14/31) shown in Table 2 with p = 1. Among those achieving viral suppression, the decline in VL was −1.24 × 10^4^ for the TM group and −3.34 × 10^4^ for the F2F subgroup. The difference in the mean decline in both groups was not statistically significant by the Snedecor Statistics (p = 1).

## DISCUSSION

In this study, we found HIV clinical indicators to be comparable between patients receiving specialty care through TM in rural Georgia and those receiving F2F care in a tertiary care center. Moreover, the mean CD4 count and the rate of increase was statistically higher in the TM group. The changes in VL and viral suppression rates were not statistically different in the study groups.

The results of this study are consistent with previous studies that have examined HIV outcomes when care is delivered through telemedicine by HIV trained physicians as compared to the F2F clinic (8,23). The mean CD4 count was statistically higher in the TM group, but the mean changes in CD4 count was similar in both groups. The changes in VL were not statistically different in both study groups.

In both clinics included in this study, some patients traveled 2-3 hours to attend the clinic. A significant proportion of low-income patients are dependent on grant funded services for HIV care coverage in rural US communities. Many residents of rural areas find primary care more accessible in their locality than specialist care. Thus, they would otherwise have significant challenges accessing HIV care from Infectious disease physicians such as what the Augusta University HIV program provides through this collaboration (15). The incidence rates of HIV in the Southern US states have been increasing despite a nationwide decline. (1) Previous studies have shown that poor access to care in remote and rural areas impacts the overall epidemiology of many “chronic” diseases. HIV patients in rural areas are more likely to be diagnosed with advance disease and have higher mortality rates. In addition, these same patients have more difficulty in finding access to care and have lower retention rates during care (13,14,24,25). Transportation constraints and the distance to the nearest available services have been consistently documented as a significant barrier to care in People Living With HIV (PLWH) (10,11,16,26). The shortage of physicians, especially the shortage of HIV trained specialists also contributes to this disparity in access to health care in rural areas (11,27). The unique telemedicine model makes it easier for physicians and other HIV trained providers to deliver the same quality of care without the restrictions of geography, travel, or time. Previous studies also demonstrated effectiveness, high patient satisfaction with telemedicine including veteran affairs patients and in PLWH. This is especially important since prior studies have demonstrated higher rates of HIV control in cohorts managed by HIV trained specialist providers (8,11,22,28).

The use of telemedicine for long term care of many of the chronic diseases including HIV care can be a particularly useful resource in these physician deprived areas. Expansion of telemedicine services to rural areas particularly in the southern United States will provide access to specialty HIV care with associated optimal viral suppression rates and a greater reduction in transmission rates, thus reducing the incidence of new cases.

An important strength of our analysis is that it was based on data obtained from routine care delivery in a rural population, and is therefore more likely to have a higher level of external validity in the South-Eastern United States than the few studies that have examined this topic in similar settings. Our findings should however be interpreted in the context of key constraints.

The observational design of our study implies that the role of residual confounding and selection bias as an explanation for our findings cannot be completely ruled out. We applied stratification and matching techniques to limit the impact of these constraints, however more rigorous prospective evaluations are warranted. In addition, the quality of the primary care services provided in our TM site though satisfactory, it might not be consistent with the quality of HIV primary care provided outside South-Eastern US. Through the Georgia Department of Public Health, our telemedicine site is a collaboration with a county health department clinic that offers primary care services and an on-site interdisciplinary team-based HIV care. The infectious disease physicians on this team are from Augusta University, providing the only telemedicine component. It is plausible that the high quality of the other components of care in the TM site accounts for the good outcomes observed.

## Conclusion

Telemedicine is as effective as face to face outpatient model for specialist delivery of HIV care in rural Georgia. Further studies should investigate relative effectiveness and acceptability of different modalities of delivering telemedicine care using rigorous prospective study designs.

To achieve eradication of HIV and control world-wide, we need to consider increasing the use of tele-medicine outreach programs in locations where specialist care is scarce or absent. This type of program can help us improve the diagnosis, management, and prevention of HIV, especially in rural areas.

## Data Availability

No external datasets in online repositories.

## Funding

This work was not supported by any funding.

## Conflicts of Interest

1. Folake J. Lawal: No conflict
2. Moshood O. Omotayo: No conflict
3. Tae Jin Lee: No conflict
4. Arni S.R. Srinivasa Rao: No conflict
5. Jose A. Vazquez: No conflict

## Notes

### Competing Interest Statement

The authors have declared no competing interest.

### Funding Statement

No funding received for this project.

### Author Declarations

Institutional Review Board - Augusta University

